# Out-of-hospital cardiac arrest incidence and temporal trends in Ecuador: a national cross-sectional study

**DOI:** 10.1101/2025.08.06.25333177

**Authors:** Ricardo S. Pinto-Villalba, Mónica Martínez, Carlos Guerrero-Calderón, Jefferson Reinoso, Rene M Abarca-Tenemasa

## Abstract

**Introduction:** Out-of-hospital cardiac arrest (OHCA) is a leading global cause of mortality, with wide regional disparities in incidence. While high-income countries have national OHCA registries, data remain scarce in Latin America, limiting the development of context-specific emergency response strategies. This study aimed to determine the national incidence and spatiotemporal distribution of OHCA in Ecuador between 2022 and 2024.

**Methods:** A retrospective observational study was conducted following STROBE guidelines. All OHCA cases reported through the Integrated Security Service (SIS ECU 9-1-1) from January 1, 2022, to December 31, 2024, were included. Temporal and geographic trends were analyzed using frequency distributions, population-adjusted incidence rates, and spatial visualization tools. Statistical analysis was performed using R; spatial data were mapped using SCImago Graphica.

**Results:** A total of 9,942 OHCA cases were recorded among 936,784 emergency events (1.06%). Annual case counts decreased from 3,635 in 2022 to 2,969 in 2024, with incidence rates falling from 20.51 to 16.52 per 100,000 inhabitants. OHCA cases exhibited seasonal peaks in January and December. Provinces such as Loja, Imbabura, and Carchi showed the highest incidence rates, while Pichincha recorded the greatest absolute number of cases. Although most provinces showed declining trends, others—such as Los Ríos— experienced increases. Geographic disparities suggest potential influences from demographic, environmental, or systemic healthcare factors.

**Conclusions:** This study provides the first national epidemiological profile of OHCA in Ecuador, revealing clear temporal and geographic variability. The findings serve as a critical step toward building an evidence base for OHCA in Ecuador and Latin America, highlighting the need for further research and the establishment of a national OHCA registry to inform policy and policy development.

**What is Known; What the Study Adds:** *What is Known:*

- Out-of-hospital cardiac arrest (OHCA) is a leading cause of mortality worldwide, with significant variation in incidence across countries.
- Most epidemiological data on OHCA originate from high-income countries with established national registries.
- Latin America lacks comprehensive national-level data on OHCA, limiting regional understanding and evidence-based health policy design.

*What the Study Adds:*

- This study provides the first national epidemiological analysis of OHCA incidence and spatiotemporal trends in Ecuador, addressing a major regional data gap.
- The findings establish a foundational dataset for Latin America that can inform future research, guide EMS system strengthening, and support the development of context-specific OHCA prevention and response strategies.

## Introduction

Out-of-hospital cardiac arrest (OHCA) represents a major global public health concern, with significant variations in incidence across countries. In the United States, data from 2022 indicate that OHCA affects approximately 356,000 individuals annually, with an estimated incidence of 88.8 cases per 100,000 inhabitants ^1,2^. In contrast, data from the same year in Spain, derived from the Out-of-Hospital Spanish Cardiac Arrest Registry (OHSCAR), report a substantially lower incidence of 24.2 cases per 100,000 inhabitants ^3^. These differences underscore the wide variability in OHCA occurrence between nations, a trend that has evolved significantly since 2010, when the incidence was reported at 54.6 and 35.5 per 100,000 inhabitants in the United States and Spain, respectively ^4^.

The availability of robust national registries has been instrumental in tracking these epidemiological trends. Such registries enable public health authorities to analyze the underlying risk factors and etiologies of OHCA, monitor changes in health system performance, and design or refine policies aimed at improving cardiovascular health and emergency response systems ^2,5^.

Trend analysis in OHCA incidence is a critical tool for health policymakers. Given that OHCA remains a leading cause of death globally – particularly as a manifestation of cardiovascular disease-comparing its trajectory with that of other conditions and sociodemographic variables allows health systems to anticipate its burden and implement measures to enhance survival outcomes ^6^.

However, unlike high-income countries in North America and Europe, many low-and middle-income countries (LMICs) lack comprehensive national registries for OHCA. This absence of data impedes accurate assessment of the condition’s local impact and often results in the adoption of public health strategies based on the epidemiological realities of high-income countries, which may not be appropriate or effective in resource-constrained settings.

Epidemiological, clinical, and demographic profiles differ widely between countries – and even more so between continents. This further complicates the applicability of foreign data to LMICs. Limited data from LMICs such as Cameroon, India, and Iran suggest OHCA incidence rates ranging from 19.9 to 190 cases per 100,000 person-years, reflecting both the diversity and lack of standardized surveillance across these regions^7^.

While it has been common practice for LMICs to base their national cardiac arrest strategies on data from high-income nations, there is a growing consensus that regional policies should be informed by local or demographically comparable evidence. In Latin America there is a lack of data around OHCA, for instance, one of the few data points comes from the city of Medellín, Colombia, where OHCA incidence was reported at 28.1 and 26.9 per 100,000 inhabitants in 2018 and 2019, respectively ^8^. Yet, these findings are limited to a single urban center and cannot be generalized nationally.

Given the critical gap in national-level data on OHCA in Latin America, the present study aims to determine the incidence of out-of-hospital cardiorespiratory arrest in Ecuador and to analyze its temporal trends over the study period. Establishing a foundational epidemiological baseline will be key for informing future policy and improving the chain of survival in the region.

## Methods

### Study Desing and Settings

In accordance with the STROBE (Strengthening the Reporting of Observational Studies in Epidemiology) guidelines ^9^, we conducted a retrospective observational study using emergency report data collected by the Integrated Security Service SIS ECU 9-1-1 between January 1, 2022, and December 31, 2024.

Ecuador, located in the northwestern region of South America, exhibits a demographic and epidemiological profile in which ischemic heart disease is the leading cause of death, accounting for 15.2% of all deaths reported in 2023. With a population of approximately 18 million, the country is experiencing a notable epidemiological transition, characterized by progressive aging (8% over 65 years of age) and increasing urbanization (73%). These factors contribute to the growing prevalence of chronic conditions such as hypertension (23%), diabetes (6.4%), and obesity (35%), according to official health data. These conditions, coupled with a high prevalence of sedentary behavior (52% reporting physical inactivity) and smoking (12%), significantly elevate the risk of cardiorespiratory arrest– a common complication of ischemic heart disease, which resulted in 13,318 deaths in 2023, with a higher incidence in men (56.39%) than in women (43.61%) ^10,11^.

In Ecuador, SIS ECU 9-1-1 is the national entity responsible for receiving and managing all emergency calls related to health, security, and disasters. When a call is made to the unified emergency number 9-1-1, it is evaluated and routed to the appropriate dispatch console based on the nature of the incident. From this platform, coordination of response units is carried out, and a service record is subsequently generated and stored in the national emergency database.

In health-related emergencies, once the primary cause of the call is identified, the prehospital evaluator determines the severity level and coordinates the deployment of either Basic Life Support (BLS) or Advanced Life Support (ALS) ambulance units. These units are responsible for providing prehospital care and transporting the patient to the appropriate medical facility. According to Ecuadorian regulations, prehospital evaluators are licensed healthcare professionals trained in remote emergency assessment ^12^. BLS units are staffed by a driver and a paramedic, whereas ALS ambulances include a driver and two paramedics, ensuring an efficient response in high-complexity scenarios ^13^.

### Study Population and Variables

The emergency report database from SIS ECU 9-1-1 serves as an official, publicly accessible source that centralizes all incidents reported across the country. For this study, we extracted data recorded between January 1, 2022, and December 31, 2024. As an initial exclusion criterion, all incidents related to security or traffic accidents were removed, allowing the analysis to focus exclusively on health-related events.

Within the medical emergency classification in the database, incidents are categorized as clinical, traumatic, psychiatric, obstetric-gynecological, or cardiorespiratory arrest, among others. For the purpose of this study, all cases of out-of-hospital cardiorespiratory arrest (OHCA) were included, provided that complete data were available, and no fields were missing. Due to the structure of the dataset, demographic information such as patient age or sex was not included and therefore could not be analyzed.

All available data were exported to an Excel spreadsheet, where they were tabulated and processed. OHCA records were organized according to their temporal (day, month, year) and geographic (province) distribution, as well as incidence frequency. This structuring enabled a detailed analysis of the spatial and temporal variability of events, facilitating the identification of potential epidemiological patterns and seasonal trends in cardiorespiratory arrest occurrences.

To ensure the quality and validity of the dataset, a three-stage review process was implemented. Three researchers – two with master’s degrees in public health and the principal investigator – independently analyzed the data to ensure consistency and accuracy. In the event of discrepancies, an inter-observer reconciliation procedure was applied, wherein inconsistencies were discussed until consensus was achieved.

### Statistical Analysis

Descriptive statistics were used to present the results, employing frequency measures and annual incidence rates of out-of-hospital cardiorespiratory arrest (OHCA). Temporal trends were visualized using line charts to illustrate fluctuations and tendencies over the study period.

Statistical analysis was conducted using R Statistical Software version 4.5.0, which allowed for the calculation of frequency distributions, standardized incidence rates and heat maps. Data visualization was performed with SCImago Graphica Beta 1.0.48, a specialized tool for scientific data representation, ensuring precise and transparent presentation of the findings.

To strengthen the methodological robustness of the study, quality control measures were implemented during data tabulation and analysis, minimizing potential interpretation biases. Additionally, the consistency of observed trends was evaluated, supporting the identification of potential seasonal or regional patterns in OHCA incidence.

In accordance with international and local ethical guidelines and given the public and anonymized nature of the dataset, this study was not subject to institutional review board (IRB) approval ^14^.

## Results

Over the course of the study period, the Integrated Security Service SIS ECU 9-1-1 recorded a total of 936,784 emergency events, of which 1.06% (n = 9,942) were classified as out-of-hospital cardiorespiratory arrests (OHCA). The year 2022 accounted for 36.56% (n = 3,635) of all reported OHCA cases, with January being the month with the highest single-month incidence across the entire period, totaling 392 cases.

During the study, the incidence of OHCA showed a declining trend between 2022 and 2024. Annual case counts fell from 3,635 in 2022 to 3,338 in 2023 (−8.2 %) and again to 2,969 in 2024 (−11.0 % versus 2023), indicating a progressive decline over the study period (**Figure 1**). For further analysis, we adjusted the data in function of the national reported population by year and location according to *Instituto Nacional de Estadística y Censos* (INEC) reports ^15^. The year-adjusted incidence rates of OHCA per 100,000 inhabitants were 20.51 in 2022, 18.71 in 2023, and 16.52 in 2024, indicating a progressive year-on-year decrease in reported OHCA events.

**Figure 1.**
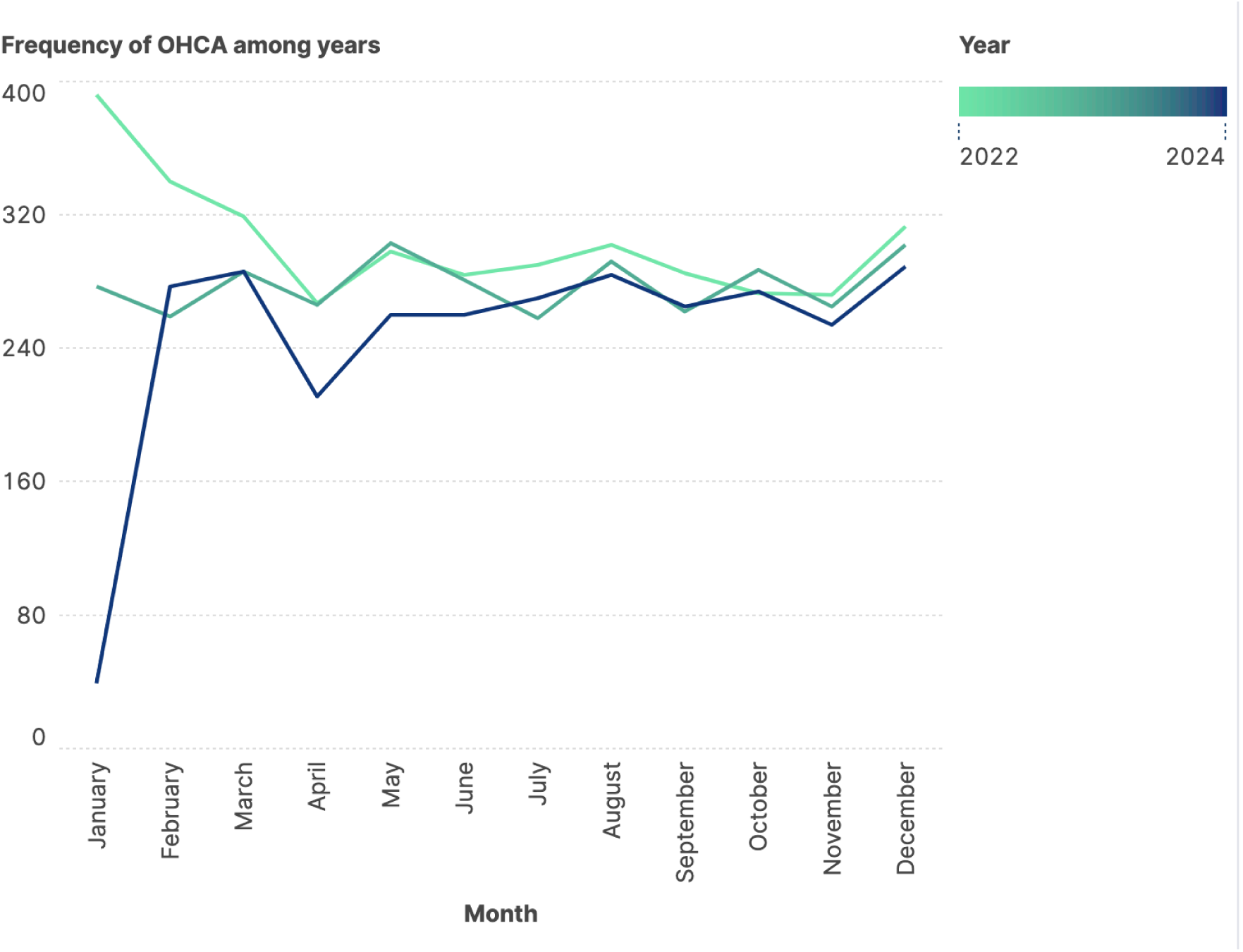
Temporal trends of OHCA in Ecuador

Monthly distributions showed a reproducible but modest seasonality (**Table 1**). In 2022 the incidence ranged from 7.3 % (April) to 10.8 % (January) of the yearly total, with values clustering tightly between 8 % and 10 % for most months. The 2023 pattern was similarly compact (7.7 %–9.1 %), with subtle peaks in May, August, and December. By contrast, 2024 exhibited an anomalously low count in January (39 cases; 1.3 %), after which frequencies returned to the typical 8 %–10 % range and again crested in December (289 cases; 9.7 %).

**Table 1.** Frequency of OHCA in Ecuador per year.

|  | Year<br>Month | 2022 |  |  | 2023 |  |  | 2024 |  |  |
| --- | --- | --- | --- | --- | --- | --- | --- | --- | --- | --- |
|  |  | Frequency | %<br>Total | %<br>Accumulated | Frequency | %<br>Total | %<br>Accumulated | Frequency | %<br>Total | %<br>Accumulated |
| Out-of-hospital<br>Cardiac<br>Arrest | January | 392 | 10.8 % | 10.8 % | 277 | 8.3 % | 8.3 % | 39 | 1.3 % | 1.3 % |
|  | February | 340 | 9.4 % | 20.1 % | 259 | 7.8 % | 16.1 % | 277 | 9.3 % | 10.6 % |
|  | March | 319 | 8.8 % | 28.9 % | 286 | 8.6 % | 24.6 % | 286 | 9.6 % | 20.3 % |
|  | April | 267 | 7.3 % | 36.3 % | 266 | 8.0 % | 32.6 % | 211 | 7.1 % | 27.4 % |
|  | May | 298 | 8.2 % | 44.5 % | 303 | 9.1 % | 41.7 % | 260 | 8.8 % | 36.1 % |
|  | June | 284 | 7.8 % | 52.3 % | 281 | 8.4 % | 50.1 % | 260 | 8.8 % | 44.9 % |
|  | July | 290 | 8.0 % | 60.2 % | 258 | 7.7 % | 57.8 % | 270 | 9.1 % | 54.0 % |
|  | August | 302 | 8.3 % | 68.6 % | 292 | 8.7 % | 66.6 % | 284 | 9.6 % | 63.6 % |
|  | September | 285 | 7.8 % | 76.4 % | 262 | 7.8 % | 74.4 % | 265 | 8.9 % | 72.5 % |
|  | October | 273 | 7.5 % | 83.9 % | 287 | 8.6 % | 83.0 % | 274 | 9.2 % | 81.7 % |
|  | November | 272 | 7.5 % | 91.4 % | 265 | 7.9 % | 91.0 % | 254 | 8.6 % | 90.3 % |
|  | December | 313 | 8.6 % | 100.0 % | 302 | 9.0 % | 100.0 % | 289 | 9.7 % | 100.0 % |

Cumulative data (**Table 1**), reveal that in all three years the mid-point of the annual burden (>50 %) was reached between June and July. However, an additional assessment of distributional assumptions with the Shapiro–Wilk test showed that the yearly aggregates were not all normally distributed: 2022 (p = 0.044) and 2024 (p < 0.001) deviated from normality, whereas 2023 did not (p = 0.271). Regardless of inter-annual variability, December consistently ranked among the three highest-incidence months, marking it as a critical period for emergency preparedness. The sharp January 2024 trough warrants further investigation to distinguish true epidemiological change from under-reporting or data-entry lag.

In a subsequent analysis the geographical distribution of out-of-hospital cardiorespiratory arrest (OHCA) cases in Ecuador revealed significant variability across provinces and over time (**Figure 2**). In 2022, the highest incidence rates per 100,000 inhabitants were observed in Loja (44.74), Imbabura (43.01), and Carchi (40.93). These provinces consistently reported elevated incidence rates across all three years, though a progressive decline was noted in each, mirroring the overall national trend (**Table 2**).

**Figure 2.**
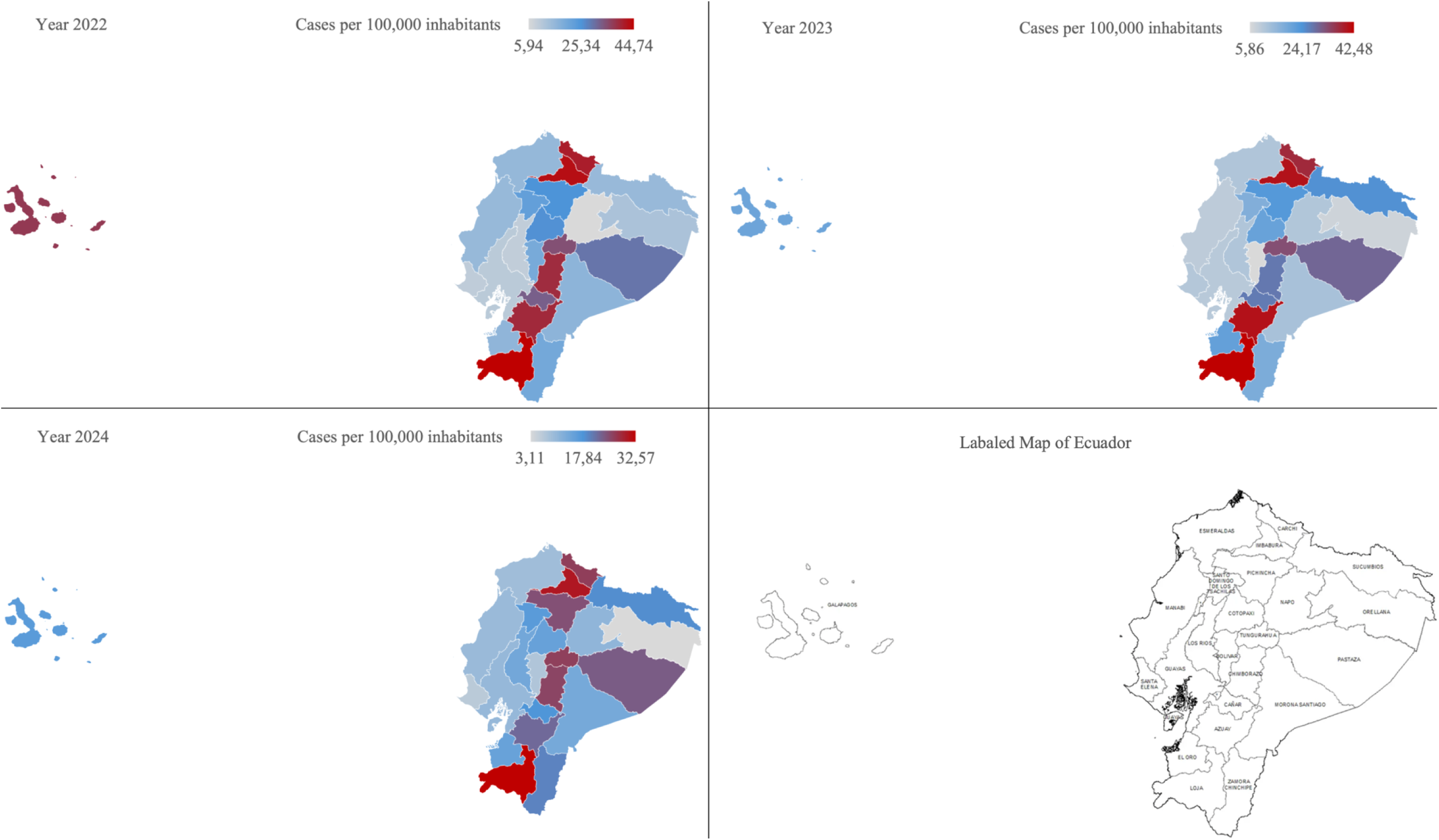
Heat map of temporal and geographical trends of OHCA in Ecuador

**Table 2.** Geographical distribution of OHCA among years.

| Year | 2022 |  |  |  | 2023 |  |  |  | 2024 |  |  |  |
| --- | --- | --- | --- | --- | --- | --- | --- | --- | --- | --- | --- | --- |
| Province | Frequency | Population | Incidence<br>rate<br>/100000 | %<br>Total | Frequency | Population | Incidence<br>rate<br>/100000 | %<br>Total | Frequency | Population | Incidence<br>rate<br>/100000 | %<br>Total |
| Azuay | 326 | 826.246 | 39,46 | 9,0 % | 334 | 828.172 | 40,33 | 10,0 % | 184 | 831.220 | 22,14 | 6,2 % |
| Bolivar | 40 | 205.544 | 19,46 | 1,1 % | 12 | 204.819 | 5,86 | 0,4 % | 15 | 204.410 | 7,34 | 0,5 % |
| Cañar | 77 | 237.584 | 32,41 | 2,1 % | 65 | 237.527 | 27,37 | 1,9 % | 42 | 237.470 | 17,69 | 1,4 % |
| Carchi | 74 | 180.776 | 40,93 | 2,0 % | 67 | 179.142 | 37,40 | 2,0 % | 47 | 177.813 | 26,43 | 1,6 % |
| Chimborazo | 193 | 493.084 | 39,14 | 5,3 % | 136 | 490.774 | 27,71 | 4,1 % | 126 | 488.857 | 25,77 | 4,2 % |
| Cotopaxi | 124 | 480.270 | 25,82 | 3,4 % | 100 | 484.792 | 20,63 | 3,0 % | 74 | 489.147 | 15,13 | 2,5 % |
| El Oro | 120 | 741.473 | 16,18 | 3,3 % | 158 | 744.811 | 21,21 | 4,7 % | 113 | 748.627 | 15,09 | 3,8 % |
| Esmeraldas | 92 | 595.674 | 15,44 | 2,5 % | 67 | 598.430 | 11,20 | 2,0 % | 55 | 601.626 | 9,14 | 1,9 % |
| Galapagos | 11 | 29.463 | 37,33 | 0,3 % | 6 | 29.808 | 20,13 | 0,2 % | 5 | 30.082 | 16,62 | 0,2 % |
| Guayas | 480 | 4.643.432 | 10,34 | 13,2 % | 478 | 4.690.613 | 10,19 | 14,3 % | 485 | 4.739.771 | 10,23 | 16,3 % |
| Imbabura | 209 | 485.913 | 43,01 | 5,7 % | 197 | 489.961 | 40,21 | 5,9 % | 150 | 494.035 | 30,36 | 5,1 % |
| Loja | 222 | 496.194 | 44,74 | 6,1 % | 211 | 496.685 | 42,48 | 6,3 % | 162 | 497.438 | 32,57 | 5,5 % |
| Los Rios | 89 | 953.060 | 9,34 | 2,4 % | 108 | 959.805 | 11,25 | 3,2 % | 135 | 968.660 | 13,94 | 4,5 % |
| Manabi | 253 | 1.670.700 | 15,14 | 7,0 % | 160 | 1.684.317 | 9,50 | 4,8 % | 166 | 1.699.434 | 9,77 | 5,6 % |
| Morona Santiago | 33 | 199.453 | 16,55 | 0,9 % | 25 | 202.851 | 12,32 | 0,7 % | 28 | 206.327 | 13,57 | 0,9 % |
| Napo | 8 | 134.768 | 5,94 | 0,2 % | 15 | 136.622 | 10,98 | 0,4 % | 15 | 138.541 | 10,83 | 0,5 % |
| Orellana | 23 | 186.166 | 12,35 | 0,6 % | 14 | 189.568 | 7,39 | 0,4 % | 6 | 192.831 | 3,11 | 0,2 % |
| Pastaza | 34 | 114.969 | 29,57 | 0,9 % | 35 | 117.062 | 29,90 | 1,0 % | 28 | 119.032 | 23,52 | 0,9 % |
| Pichincha | 821 | 3.231.547 | 25,41 | 22,6 % | 754 | 3.250.537 | 23,20 | 22,6 % | 807 | 3.272.265 | 24,66 | 27,2 % |
| Santa Elena | 38 | 399.185 | 9,52 | 1,0 % | 43 | 400.829 | 10,73 | 1,3 % | 26 | 403.478 | 6,44 | 0,9 % |
| Santo Domingo de los Tsachilas | 116 | 513.692 | 22,58 | 3,2 % | 90 | 518.576 | 17,36 | 2,7 % | 87 | 523.524 | 16,62 | 2,9 % |
| Sucumbios | 31 | 207.221 | 14,96 | 0,9 % | 51 | 206.360 | 24,71 | 1,5 % | 38 | 205.253 | 18,51 | 1,3 % |
| Tungurahua | 197 | 573.786 | 34,33 | 5,4 % | 189 | 576.407 | 32,79 | 5,7 % | 152 | 579.082 | 26,25 | 5,1 % |
| Zamora Chinchipe | 23 | 115.101 | 19,98 | 0,6 % | 21 | 116.363 | 18,05 | 0,6 % | 23 | 117.650 | 19,55 | 0,8 % |
| Undelimited area | 1 |  |  |  | 2 |  |  | 0,1 % | 0 |  |  | 0,00 % |

In absolute terms, the province of Pichincha consistently reported the highest number of OHCA cases throughout the study period, accounting for 22.6% in 2022, 22.6% in 2023, and 27.2% in 2024. However, when adjusted for population, its incidence rates remained moderate, ranging from 23.20 to 25.41 per 100,000 (**Table 2**). Conversely, Guayas, despite being the most populous province, exhibited one of the lowest adjusted incidence rates (around 10.2 per 100,000 across all three years), suggesting a relatively lower risk profile or possible underreporting.

Temporal trends indicate a generalized reduction in OHCA incidence across most provinces from 2022 to 2024 (**Table 2**). For example, Azuay reported a drop from 39.46 per 100,000 in 2022 to 22.14 in 2024, and Tungurahua from 34.33 to 26.25, highlighting a downward trend in both frequency and rate. Some provinces, such as Los Ríos, deviated from this pattern, showing a year-on-year increase in incidence—from 9.34 in 2022 to 13.94 in 2024, warranting further investigation.

Provincial contributions to the total national OHCA burden remained relatively stable for most territories. However, smaller provinces such as Galápagos, Napo, and Zamora Chinchipe reported low case numbers and population-adjusted rates, contributing under 1% of national cases annually.

These findings underscore both spatial and temporal disparities in OHCA incidence in Ecuador, with implications for the allocation of prehospital resources, targeted prevention strategies, and local health system strengthening, particularly in provinces with persistent or rising incidence trends.

## Discussion

This study presents the first national-level epidemiological description of out-of-hospital cardiorespiratory arrest (OHCA) in Ecuador, contributing to the limited body of data available from Latin America. Our findings reveal a progressive decline in OHCA incidence over a three-year period, a modest seasonal pattern, and notable geographical disparities across the country. These results provide a valuable reference point for future policy development, regional benchmarking, and hypothesis generation.

The observed decline in national OHCA incidence, from 20.5 to 16.5 cases per 100,000 inhabitants between 2022 and 2024, aligns with reports from several high-income countries where incidence rates have plateaued or declined following the implementation of cardiovascular prevention strategies and system-level improvements in emergency care ^16–18^. While causality cannot be established in our study, this trend may reflect the effects of improved risk factor management (e.g., hypertension, dyslipidemia, tobacco cessation), public health interventions, and better access to treatment – hypotheses supported by evidence from large-scale cohort studies ^19,20^.

The seasonal distribution of cases, with consistent peaks in December and January, is also consistent with international literature reporting higher cardiac arrest incidence during winter months in temperate climates, likely associated with cold exposure, respiratory infections, and increased psychosocial stress ^21^. Although Ecuador’s equatorial climate lacks marked seasonal variation, the observed peaks may be influenced by behavioral changes related to year-end festivities, including increased alcohol intake, reduced physical activity, and dietary excess, as well as possible influenza-like illness circulation, may underlie these variations–consistent with literature linking influenza peaks to elevated cardiovascular events ^22^.

The geographical disparities in incidence across Ecuador are particularly notable. Provinces such as Loja, Imbabura, and Carchi consistently reported the highest adjusted incidence rates throughout the study period. These areas share common geographical characteristics such as high altitude (>2 000 m above sea level), rural dispersion, and limited EMS coverage. This results contrast whit previous studies which have identified high-altitude exposure as a possible protective risk factor for cardiovascular complications due to genetic adaptive patterns in highlanders ^23,24^. Moreover, rural and highland areas may experience longer EMS response times, further reducing the likelihood of successful resuscitation, as reported in previous studies ^25^.

In contrast, Guayas, Ecuador’s most populous province, reported one of the lowest adjusted OHCA incidence rates, despite having the highest population density. This finding could reflect better healthcare infrastructure and urban EMS coverage, although the possibility of underreporting or differences in data collection practices cannot be excluded. Interestingly, Pichincha consistently accounted for the highest absolute number of cases but maintained only moderate adjusted incidence rates – consistent with findings in other urban centers where absolute burden correlates with population size, but adjusted rates are influenced by access to care and public health resources ^26,27^.

The increasing trend observed in Los Ríos – from 9.3 to 13.9 cases per 100,000 between 2022 and 2024 – diverges from the national pattern and raises questions about local factors influencing risk. Similar upward trends in specific regions have been linked to rapid urbanization, demographic shifts, and reduced access to primary care ^28,29^.

### Limitations

While these findings provide novel insights, they must be interpreted within the limitations of the study. The use of an administrative database without patient-level demographic, clinical, or outcome data precludes analysis of survival rates, risk stratification, or age-adjusted comparisons. Furthermore, underreporting or misclassification of cases may have occurred, particularly in remote or underserved areas. Despite these limitations, the consistency of trends across multiple provinces strengthens the internal validity of the results and allows for the generation of several research hypotheses.

### Implications for practice and future research

Future efforts should focus on the development of a dedicated national OHCA registry, aligned with Utstein-style reporting standards, to enable monitoring of outcomes, identification of at-risk populations, and evaluation of interventions. Public health initiatives should prioritize provinces with persistently elevated incidence, particularly those in rural and high-altitude settings. Additionally, the integration of community-based cardiopulmonary resuscitation (CPR) training, broader deployment of automated external defibrillators (AEDs), and improvements in EMS coverage may help reduce the burden of OHCA nationwide.

## Conclusion

Our findings reveal a progressive decrease in OHCA incidence in Ecuador between 2022 and 2024, accompanied by seasonal patterns and marked regional heterogeneity. The results highlight the need for targeted public health interventions, improves in emergency planning, and the establishment of a national OHCA registry to inform policy and policy development in Ecuador and comparable Latin American contexts.

## Data Availability

All data will be available upon request

## Acknowledgments

None

## Contributors

### CRediT Author Statement

Pinto-Villalba Ricardo S.: Conceptualization, Methodology, Software, Validation, Formal Analysis, Investigation, Resources, Data Curation, Supervision, Project Administration, Visualization, Writing – Original Draft, Writing – Review & Editing

Martínez Mónica: Conceptualization, Validation, Formal Analysis, Investigation, Writing – Original Draft, Writing – Review & Editing

Guerrero-Calderón Carlos: Conceptualization, Software, Investigation, Visualization, Writing – Original Draft, Writing – Review & Editing

Reinoso Jefferson: Conceptualization, Investigation, Writing – Original Draft, Writing – Review & Editing

Abarca-Tenemasa Rene M.: Conceptualization, Validation, Investigation, Resources, Supervision, Project Administration, Visualization, Writing – Original Draft, Writing – Review & Editing.

